# Mixed method estimation of population level HIV viral suppression rate in the Western Cape, South Africa

**DOI:** 10.1101/2020.03.19.20038745

**Authors:** Elton Mukonda, Nei-Yuan Hsiao, Lara Vojnov, Landon Myer, Maia Lesosky

## Abstract

**Introduction:** There are few population-wide data on viral suppression (VS) that can be used to monitor programmatic targets in sub-Saharan Africa. We describe how routinely collected viral load (VL) data from ART programmes can be extrapolated to estimate population VS and validate this using a combination of empiric and model-based estimates.

**Methods:** VL test results from were matched using a record linkage algorithm to obtain linked results for individuals. Test- and individual-level VS rates were based on test VL values <1000 cps/ml, and individual VL <1000 cps/mL in a calendar year, respectively. We calculated population VS among people living with HIV (PLWH) in the province by combining census-derived mid-year population estimates, HIV prevalence estimates and individual level VS estimates from routine VL data.

**Results:** Approximately 1.9 million VL test results between 2008 – 2018 were analysed. Among individuals in care, VS increased from 85.5% in 2008 to 90% in 2018. Population VS among all PLWH in the province increased from 12.2% in 2008 to 51.0% in 2017. The estimates derived from this method are comparable to those from other published studies. Sensitivity analyses showed that the results are robust to variations in linkage method, but sensitive to the extreme combinations of assumed ART coverage and population HIV prevalence.

**Conclusion:** While validation of this method in other settings is required, this approach provides a simple, robust method for estimating population VS using routine data from ART services that can be employed by national programmes in high-burden settings.

## Introduction

Viral load (VL) monitoring is the current standard for evaluating treatment effectiveness and transmission risk in people living with HIV (PLWH). Subsequent to World Health Organisation (WHO) guidelines supporting the implementation of VL monitoring in 2013 for all HIV-infected adults on ART [1], there has been a rapid roll-out and scale-up of VL monitoring in sub-Saharan Africa [2]. Programmatic evaluation of public sector monitoring requires systematic, and often digital, data collection, storage and analysis. Most African countries have only recently developed capacity for digital collection and storage of routine laboratory data [3,4], though others, such as Kenya (https://viralload.nascop.org/) and Uganda (https://vldash.cphluganda.org/) have embraced digital and open dashboards as one part of reporting. Routine data represents a rich source of program evaluation which is often overlooked due to concerns regarding completeness, timeliness, representativeness and accuracy [5]. This ultimately hampers efforts to evaluate micro- and macro-level programme interventions or policy changes, for example, evaluating treatment effectiveness and transmission risk in people living with HIV (PLWH). Commonly, despite the high cost and low frequency, there is a reliance on intermittent community surveys and multiple indicator cluster surveys (MCIS) to evaluate micro and macro program interventions [5].

In parallel, UNAIDS ‘90-90-90’ targets [6] require the use of VL monitoring to assess the ‘third-90’. Meeting the ‘90-90-90’ targets requires an approximate population viral suppression rate among all PLWH, including those not in care, of approximately 73%. Most of the published work evaluating the ‘90-90-90’ targets have relied on special surveys [7-10] or mathematical models [11,12], and when routine data are used, the results typically provide only a snapshot of the progress for a 1-year period [13,14].

In this study we develop a method for the evaluation of progress towards the ‘third 90’ and the overall ‘90-90-90’ target in the Western Cape, South Africa using routinely collected public sector laboratory data, regional population census estimates and model-based HIV prevalence estimates. We demonstrate a novel mixed method approach, as well as a novel record linkage algorithm to estimate age and sex stratified viral suppression rates among all PLWH in the Western Cape, South Africa.

## Methods

### Viral load measures

According to the South African guidelines [15], routine viral load monitoring for individuals receiving antiretroviral therapy (ART) is 6 and 12 months after ART initiation and annually thereafter. Elevated VL>1000 copies/mL triggers additional testing, per guidelines to occur within 6 months of the initial elevated VL. For each routine viral load request, a standardised request form containing patient identifiers are manually captured into the laboratory information system at the receiving laboratory before the patient sample is centrifuged and tested using two locally validated and equivalent platforms: the Abbott Realtime HIV-1 (Abbott Molecular, Des Plaines, USA) or Roche Cobas Ampliprep/Cobas TaqMan HIV-1 (Roche Diagnostics, Branchburg, USA) assays.

### Data

We reviewed routine VL test results carried out by the National Health Laboratory Service (NHLS) in the two laboratories based in the Western Cape province, South Africa between 1 January 2008 and 30 September 2018. The NHLS is the largest diagnostic pathology service in South Africa providing laboratory testing to public sector patients (over 80% of the South African population attend public sector services [4]) through a national network of laboratories. These data were initially in the form of separate unlinked individual VL test results. Minimal clinical and individual characteristics were available, including date of test, sex, age or date of birth (partial), and location of test request (health care facility). Results from facilities in other provinces, from private sector healthcare facilities, and tests taken as part of a clinical trials or research studies were excluded. The resulting data set represents a catchment area consisting of all public-sector ART clinics and hospitals servicing individuals on ART in the Western Cape province of South Africa.

### Data processing and record linkage

Due to the variable presence of reliable unique identifiers in the data a custom record linkage procedure was developed. Individuals who had complete matching identifier information (including first and second names, date of birth, and patient record identifier) were separated into the reference set. The remaining results were split into data sets based on the quality of the identifying information. Pairwise evaluations between the reference set and the comparative data sets were made and similarity scores using the Jaro-Winkler (JW) algorithm [16] and hierarchical clustering using the Jaro-Winkler distance metric were calculated for first and second names independently. Scores were combined, and any similarity score above a specified threshold was used to indicate a match. Matches were integrated into the reference set, and the algorithm run iteratively until no further matches could be identified. The same procedure was performed iteratively by using the best of the comparative datasets as the reference. Different thresholds for the distance metric and summary scores were evaluated as part of a sensitivity analyses, this and full details of the record matching algorithm can be found in the online supplement.

## Statistical methods

### Test-level viral suppression

Each VL result was classified as “suppressed” or “not suppressed”. Two definitions of viral suppression (VS) were considered: the WHO threshold of a test VL <1000 copies/mL, and VL < 400 copies/mL. The overall proportion of suppressed tests by any strata or period was calculated by taking the total number of test results below the threshold and dividing it by the total number of tests in that period.

### Person-level viral suppression

The proportion of HIV-infected individuals assumed to be in care and on treatment who were virally suppressed in each year was calculated as the total number of individuals whose last test in the year had VL <1000 copies/mL (or VL <400 copies/mL) divided by the total number of individuals having a VL test in that year. As part of a sensitivity analysis, individual level viral status was also evaluated using the first test of the year.

### Population-level viral suppression

The estimated number of PLWH in each year was calculated using data from three sources: the South African census, a mathematical model, and the routine data described in this work. End of year population estimates were interpolated from consecutive mid-year population estimates produced by Statistics South Africa [17-27] under the assumption that the population is growing at a constant rate (exponential growth model).

The annual HIV prevalence for the Western Cape province was estimated from the Thembisa model (a mathematical model of the South African HIV epidemic, accessed June 2019) [28]. The product of the annual HIV prevalence and the total number of individuals at risk (end of year adult population) is the estimated number of PLWH in the Western Cape. Age and sex strata sub-estimates were calculated in the same way, making provisions for the availability of annual age and sex specific HIV prevalence rates in the Thembisa model. The number of people in care and on treatment was estimated using the number of unique individuals with any viral load measurement during the given period. We further assumed that all individuals not in care (the difference between the number estimated to be living with HIV and the number of individuals reported in the VL database) would have VL>1000 copies/mL (or >400 copies/mL). This assumption was subject to sensitivity analyses.

Population-level viral suppression was estimated by taking the number of individuals virally suppressed in a given period and dividing by the total number of people living with HIV (both in and out of care). Considering that routine test data are only available for a fraction of 2018, population-level suppression was only estimated for the period 2008-2017. We compared our individual- and population-level viral suppression estimates with other published estimates or special surveys, and with the standard target represented by the ‘third-90’.

Estimates were tabulated by frequency (percent) and visualised using line and bar charts. 95% confidence intervals for proportions were calculated. All analyses and record linkage were done in R version 3.6 (R Core Team, Vienna, Austria). Use of data was approved by the University of Cape Town Human Research Ethics Committee and conformed to the principles embodied in the declaration of Helsinki. The need for individual consent was waived by these committees.

## Results

### Test-level suppression

A total of 1,985,883 VL tests were available from 533 sites between January 1, 2008 to September 30, 2018. Services scaled up from an average of 5,128 VL tests per month in 2008 to 27,558 per month in 2018, an increase of nearly 500%. Approximately two-thirds of the tests carried out during the study period came from women. The age distributions for males and females are similar in shape with males a median (IQR) age of 38 years (IQR: 31-44) versus 34 years (IQR: 28-40) for females (Table 1). The aggregate proportion of virally suppressed tests (VL<1000 cps/mL) was higher among women in care than men in care (85.0% vs 82.5%; p<0.001) (Table 1). Similar results were found when viral suppression was assessed at 400 copies/mL, and over each calendar year (Table 1, Supplement Table 1). Overall test-level viral suppression rates ranged from 80% to 87% during the study period (Table 2) with over 80% of test results coming from individuals between the ages of 25-54 (Supplement Table 2-3).

**Table 1:**
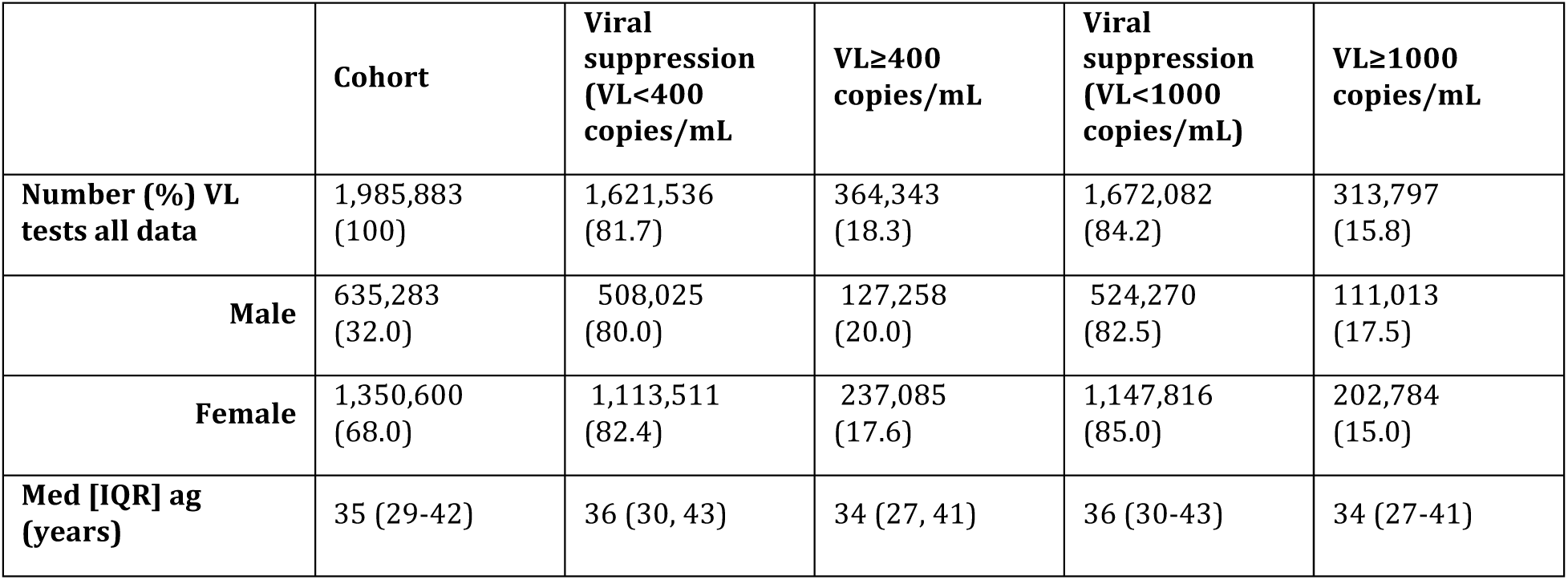
Characteristics of the viral load tests requested from NHLS by viral load level (2008-2018)

|  | **Cohort** | **Viral suppression (VL<400 copies/mL)** | **VL≥400 copies/mL** | **Viral suppression (VL<1000 copies/mL)** | **VL≥1000 copies/mL** |
| --- | --- | --- | --- | --- | --- |
| **Number (%) VL tests all data** | 1,985,883<br>(100) | 1,621,536<br>(81.7) | 364,343<br>(18.3) | 1,672,082<br>(84.2) | 313,797<br>(15.8) |
| **Male** | 635,283<br>(32.0) | 508,025<br>(80.0) | 127,258<br>(20.0) | 524,270<br>(82.5) | 111,013<br>(17.5) |
| **Female** | 1,350,600<br>(68.0) | 1,113,511<br>(82.4) | 237,085<br>(17.6) | 1,147,816<br>(85.0) | 202,784<br>(15.0) |
| **Med [IQR] ag (years)** | 35 (29-42) | 36 (30, 43) | 34 (27, 41) | 36 (30-43) | 34 (27-41) |

**Table 2:** Summary and comparison of test-level and person-level viral suppression (VL<1000 copies/mL) between 2008 and 2018 for all tests and for individuals in care, disaggregated by sex.

|  | Male |  | Females |  | Overall |  |
| --- | --- | --- | --- | --- | --- | --- |
| Year | Tests <1000<br>copies/mL N<br>(%) | Individuals on<br>ART<1000<br>copies/mL N<br>(%) | Tests <1000<br>copies/mL N<br>(%) | Individuals on<br>ART<1000<br>copies/mL N<br>(%) | Tests <1000<br>copies/mL N<br>(%) | Individuals on<br>ART<1000<br>copies/mL N<br>(%) |
| 2008 | 14557(83.5) | 10738(84.1) | 28696(85.1) | 21297(86.2) | 43253(84.6) | 32035(85.5) |
| 2009 | 26973(84.7) | 16663(84.4) | 53850(86.6) | 32789(86.7) | 80823(85.9) | 49452(85.9) |
| 2010 | 26578(83.8) | 19797(85.2) | 52017(84.6) | 38813(86.3) | 78595(84.3) | 58610(85.9) |
| 2011 | 33756(78.9) | 26907(83.7) | 67789(79.9) | 53992(85.2) | 101545(79.6) | 80899(84.7) |
| 2012 | 41971(81) | 34263(85.3) | 85985(82.9) | 69903(87.5) | 127956(82.3) | 104166(86.7) |
| 2013 | 50801(80.9) | 41138(85.6) | 104750(82.6) | 85690(87.5) | 155551(82.1) | 126828(86.9) |
| 2014 | 52128(81.1) | 44108(85.7) | 115523(83.9) | 96100(88.1) | 167651(83) | 140208(87.3) |
| 2015 | 61358(82.2) | 52614(86.5) | 142353(85.4) | 117255(89.2) | 203711(84.4) | 169869(88.3) |
| 2016 | 72922(83) | 62655(87.9) | 166176(85.7) | 135684(90) | 239098(84.9) | 198339(89.3) |
| 2017 | 79031(83.5) | 69454(88.3) | 183101(86.6) | 152080(90.7) | 262132(85.7) | 221534(89.9) |
| 2018^ | 64195(85) | 60726(88.6) | 147576(87.7) | 133768(90.7) | 211771(86.9) | 194494(90) |

^ 2018 is a partial year estimate, Jan - Sept

### Record linkage

The linkage procedure described resulted in 8 different patient-level cohorts based on the linkage method (Supplement Table 6). These cohorts were then compared to the number of people on ART as reported by the Western Cape Department of Health (DOH) [29] (Supplement Figure 1) to evaluate linkage validity. While deterministic linkage (Full name or ID) produces cohorts with higher numbers of unique individuals, the overall percentage differences from the Western Cape Department of Health estimates are higher than those produced from a combination of deterministic and probabilistic linkage. The linkage methods with Jaro-Winkler distance metric thresholds of 0.9 and 1 (perfect linkage) produced results closest to the DOH estimates. The method with Jaro-Winkler metric similarity threshold of 0.9, which accounts for approximate matches rather than exact matches only (similarity threshold of 1), was chosen as the baseline linkage scenario. This gives rise to the cohort reported for individual-level and population-level suppression with a total of 474,595 unique individuals in care and a median of 7 (IQR: 4-10) tests per individual.

### Individual-level suppression

As with test-level suppression rates, person-level suppression rates for those in care (VL<1000 copies/mL) were consistently higher for females (range: 84.5% to 90.7%) than for males (range: 83.1% to 88.6%) between 2008 and 2017, irrespective of age group (Table 2). All age and sex strata demonstrated improvement in viral suppression rates, except for children between the ages of 1 and 5 years (Figure 1) for whom viral suppression rates appear flat or potentially in decline. The lowest rates of viral suppression in the Western Cape were for children between the ages of 0-2 years with the highest rates for people older than 65 years (Figure 1, Supplement Table 2-3). However, infants in this province receive a pre-ART VL as confirmation for their PCR diagnosis, potentially impacting the VS estimate. In all strata, individual-level viral suppression was consistently higher than test-level viral suppression, pointing to the importance of record linkage. This difference may also reflect the potential for oversampling of viremic individuals due to policy guidelines (individuals with VL>1000 cps/mL should be re-tested within 3-6 months).

**Figure 1.**
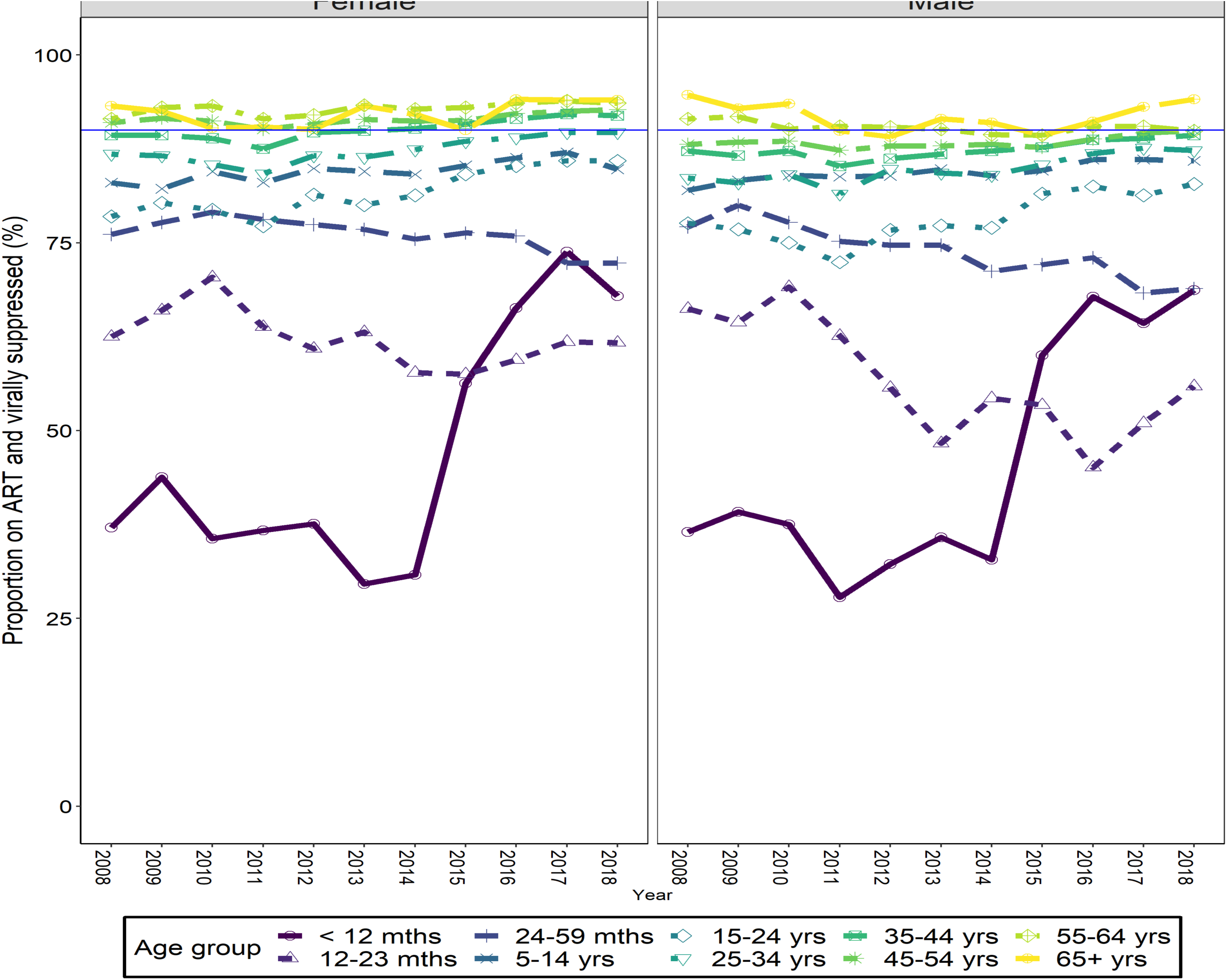
Proportions of individuals in care who are virally suppressed in the Western Cape disaggregated by age and sex.

### Population-level suppression

The estimated rate of population-level viral suppression among all PLWH, both in care in the public health sector and outside of care, increased from 12.3% in 2008 to 51% in 2017, growth of more than 300% (Table 3). At the end of 2017, an estimated 51% of PLWH were estimated to be virally suppressed, while an estimated 90% of PLWH and enrolled in public sector services were virally suppressed (Table 2,3). Our estimates varied slightly on use of VL<400 copies/mL as a threshold (Supplement Table 7). The estimates for population-level viral suppression did not vary substantially with linkage method, though there are noticeable differences when comparing deterministic linkage and the combination of deterministic and probabilistic linkage (Supplement Table 8). Disaggregating by age and sex (Supplement Figure 2) reveals that women living with HIV had consistently higher population viral suppression rates compared to men living with HIV, while youths living with HIV (ages 15-24 years) appeared to have the lowest rates of population viral suppression (range: 2.8% to 39%) in the period. Women living with HIV and between 25 and 49 appear to be making the most progress with an estimated 72% being virally suppressed in 2017.

**Table 3:** Estimated population viral suppression rate for all people living with HIV in the Western Cape

| **Year** | **End of year population<sup>^</sup>** | **Total HIV prevalence (%) <sup>#</sup>** | **HIV infected population** | **Individuals with VL<1000 copies/mL** | **Proportion virally suppressed<sup>^^</sup>** |
| --- | --- | --- | --- | --- | --- |
| 2008 | 5,308,512 | 4.9% | 261,332 | 32035 | 12.3% |
| 2009 | 5,288,519 | 5.2% | 275,266 | 49452 | 18.0% |
| 2010 | 5,254,963 | 5.5% | 287,057 | 58610 | 20.4% |
| 2011 | 5,455,552 | 5.7% | 310,433 | 80899 | 26.1% |
| 2012 | 5,819,473 | 5.9% | 342,790 | 104166 | 30.4% |
| 2013 | 6,063,550 | 6.1% | 368,182 | 126828 | 34.4% |
| 2014 | 6,157,898 | 6.2% | 383,989 | 140208 | 36.5% |
| 2015 | 6,222,410 | 6.4% | 397,189 | 169869 | 42.8% |
| 2016 | 6,396,236 | 6.5% | 416,515 | 198339 | 47.6% |
| 2017 | 6,564,961 | 6.6% | 434,634 | 221534 | 51.0% |
| 2018* | 6,655,523 | 6.7% | 446,303 | 194494 | 43.6% |

<sup>^</sup> Interpolated from SA mid-year population estimates
\*Estimates for the end of September 2018
<sup>#</sup> Obtained from the Thembisa model
<sup>^^</sup> Assumes those not in care have VL>1000 cps/mL

### Comparison to other studies

Estimates for the third-90 found in this study using routine laboratory data were similar to those produced from models [11] for the Western Cape, and those obtained from other studies [7-10] for other South African populations (Figure 2). The major differences are likely attributable to the different populations and age groups for which the other estimates were produced [8-10,13], and different implementations of guidelines by region or province. Definition of viral suppression (VL<400 copies/mL vs VL<1000 copies/mL) [11] and the fact that estimates produced in this study did not verify individuals on ART may also play a role in the variation of estimates.

**Figure 2.**
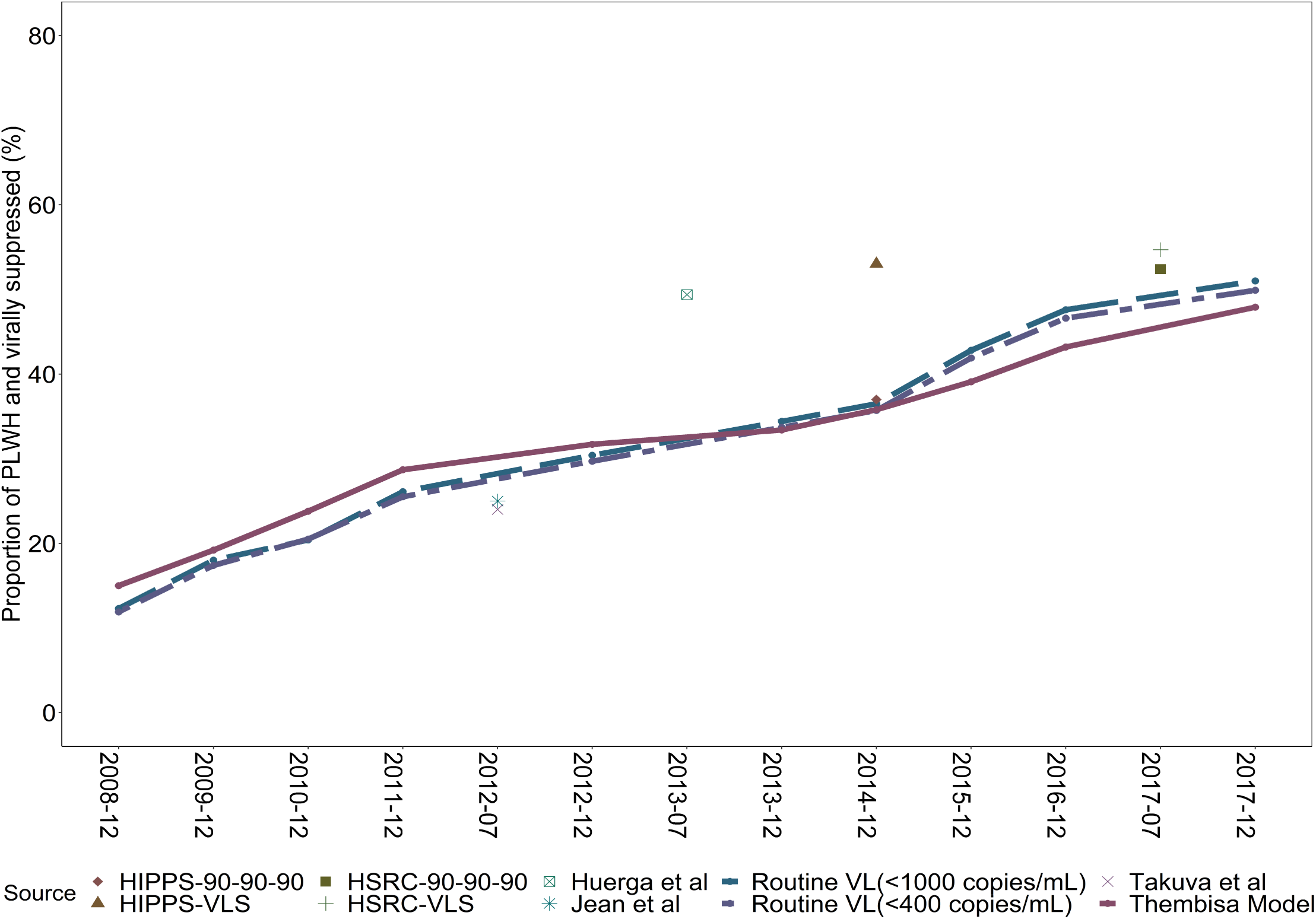
Method based estimation of population viral suppression among all people living with HIV (PLWH) in the Western Cape (dashed lines), compared to mathematical model based (purple solid line) and cross-sectional survey estimates (points) from 2008 - 2017.

## Discussion

Using routine laboratory data, we have provided valuable insight into the program achievements of the South African public sector HIV services in the Western Cape. The aim of this study was to give the year-on-year progress made towards the third 90 in the Western Cape using routinely collected data. An in-depth analysis of the progress made towards the third 90 may help to identify populations and areas which may require additional support for HIV testing, linkage, ART rollout strategies and adherence/retention [30]. The results show clear evidence of substantial improvement in test-level and estimated population-level viral suppression rates in the study period, against a background of a massive scale up of HIV testing and ART services, pointing to the effectiveness of the South African HIV treatment programmes. There remain areas of concern however, and the ‘third 90’ target appears to be only partially met. The proportion of HIV-infected individuals who are virally suppressed steadily increased between 2008 and 2018, though disaggregation by age and sex reveals stark differences between groups throughout the period.

Women in care in the Western Cape have achieved the third-90 target with 90% of women on ART virally suppressed in 2018. Women received significantly higher numbers of tests when compared to men, and had marginally higher proportions of tests with VL <1000 copies/mL. This is in line with other data from this area and other regions, where retention of men in ART services tends to be worse and men show poorer rates of maintenance of viral suppression [31]. The overrepresentation of women in HIV care services may be due to the strong maternal and child health platform, or stronger care seeking behaviour among women in the Western Cape [30,32,33] as entry and retention into these programs were better. This supports the call for further targeting of HIV testing and ART initiation and retention strategies aimed at men.

While there is an overall improvement in viral suppression rates among individuals for most of the age groups, suppression for children aged between 0 and 5 years appears to be decreasing over the period. This phenomenon is unexplained by the available data and would warrant further investigation but could be a consequence of limitations of the data, or of implementation concerns such as difficulty in drug administration or non-optimal drugs. HIV-infected individuals between 15 and 24 years had markedly low viral suppression rates and are a group of concern. This is thought to be primarily a result of lower adherence to ART in this age group, though other possible reasons include poor transition from paediatric to adult services, poor linkage to care and sub-optimal disclosure of adolescents’ HIV status [34]. Older age groups in care do better than younger age groups, again in line with other findings [35]. Only 17% of all tests were for individuals either above 55 years and under 25 years. This is in line with the low rates of HIV testing among children [35], and older individuals, who are generally not targeted for testing and HIV prevention [34]. Proportions of men and women who were virally suppressed and over the age of 55 were consistently above 90% between 2008 and 2017.

Population-level viral suppression estimates produced in this study were consistent with estimates produced from mathematical modelling [11] for the Western Cape and those produced in other studies [7-10,13,14,37]. The most recent of these surveys was the Fifth South African National HIV Prevalence, Incidence, Behaviour and Communication Survey (SABSSM V) conducted by the Human Sciences Research Council (HSRC) (37) in 2017. This study produced estimates for the ‘third-90’ and viral load suppression prevalence, a measure which is similar in calculation to population viral suppression. Estimates for population viral suppression prevalence and the ‘third-90’ for the Western Cape province from the HSRC study were 54.7% and 52.4%, respectively. Both are comparable to the estimate obtained in this study for 2017 (51.0%). Additionally, the disaggregated estimates are also consistent with the findings in this study that females had higher VL suppression and that among youth and adult PLWH, those aged between 15-24 had the lowest VL suppression, while those above 45 had the highest suppression rates.

There are a few key assumptions underlying this analysis. In particular, the assumption is that there is good VL testing coverage in the Western Cape and that population mobility (receiving care elsewhere) is not a major factor. This is reasonable to assume in this province with an extensive and low-cost to user ART provision program, but may not hold in other settings. While individuals not in care were assumed to be viraemic, individuals in care, on treatment but without access to viral load were not explicitly accounted for, nor were individuals receiving HIV care in the private sector accounted for, though they make up a very small proportion of the population living with HIV in South Africa.

This analysis is not without limitations as it primarily depends on uniquely identifying individuals in the population. The estimates of population-level suppression are therefore dependent on the robustness of the record linkage procedure. In the absence of a reliable unique identifier there is still room for erroneous matching. However, the fact that approximately 85% of the data had reliable and consistent identifiers prior to probabilistic linkage suggests that the overall estimates are likely robust. Secondarily, this analysis depends on the usage of census and model-based estimates of the population living with HIV. While population estimates were obtained and extrapolated from census data, there were no empirical data from which to extract annualised provincial HIV-prevalence rates, disaggregated by age group and sex over the period of interest. As a result, population-level suppression estimates are only as reliable as the secondary data used to produce them. The Thembisa model [11] is a reliable source of HIV prevalence estimates for the South African population, but does not extend to other settings. A possible source of annualised prevalence estimates is the Spectrum model, which is used by national programs and UNAIDS to prepare annual estimates of the status of the HIV epidemic in 160 countries [38]. An additional weakness of the study is that the estimates given do not factor in the change in policy from the bi-annual tests to annual tests circa 2010, nor the broadening of treatment eligibility that occurred over the study period [39].

Importantly, our study used routinely collected data and did not require additional or specialised surveys, leveraging an important source of data. Despite the lack of a reliable unique identifier, we demonstrated a record linkage procedure using available identifiers to assign tests to individuals in the dataset. In cases where there is no identifying information with which to perform the record linkage, test-level estimates can be used. However, care must be taken in interpretation as test-level viral suppression is likely to underestimate the person-level, and population-level viral suppression due to oversampling of individuals who are viremic, in line with most guidelines that suggest re-testing of individuals after evidence of viremia. Estimates from this study were comparable to specialised surveys carried out for the purpose of population-level viral suppression, supporting a mixed methods approach for routine data.

## Conclusions

There has been a significant scale up in the provision of VL testing in the Western Cape and South Africa more generally, along with clear evidence of improving rates of viral suppression. While it is evident that significant progress has been made towards the third 90, the target to have 73% of HIV-infected individuals on ART and virally suppressed by 2020 is unlikely to be met. Routinely collected test-level data alone should not be used for estimates of person- or population-level viral suppression rates without record linkage being performed as it consistently underestimates the rate of viral suppression, at least in settings where follow up viral load tests are undertaken in response to raised viral load. The regular use of routinely collected viral load data in conjunction with an understanding of ART coverage and population in care could, however, help bridge the gaps of knowledge and aid in identification of vulnerable populations that require concerted efforts to achieve the overall 90-90-90 targets. This approach has promise in settings where reliable population and HIV prevalence estimates are available and may allow for more frequent and less expensive program assessment.

## Data Availability

The data is owned by the South African National Health Laboratory Services and includes personal information including identifiers, therefore cannot be made publicly available.

